# Preparedness of Sub-Saharan African Countries to Address Climate Change and Health Impact: A Scoping Review

**DOI:** 10.1101/2024.11.28.24318138

**Authors:** Aminata Kilungo, Godsgift N. Chukwuonye, Victor Okpanachi, Mohamed Hussein

**Affiliations:** Department of Community, Environment and Policy, University of Arizona, Tucson, Arizona; Department of Environmental Sciences, The University of Arizona, Tucson, Arizona; Department of Environmental and Occupational Health, Muhimbili University of Health and Allied Sciences, Dar es Salaam, Tanzania

**Keywords:** Climate change, health, sub-Saharan Africa

## Abstract

Climate change severely threatens global public health, with sub-Saharan Africa (SSA) projected to experience profound impacts. This scoping review aimed to provide a comprehensive overview of current research on climate change and its health implications in SSA while identifying research gaps and outlining the necessary resources and policy interventions to strengthen public health resilience in the region. Literature was retrieved from four databases (PubMed, Scopus, Embase and Web of Science) using the keywords “climate change,” “health,” and “sub-Saharan Africa”. A total of 7851 journal articles were identified from the initial search, and after screening, 153 studies were included for review. The included studies were published between January 2001 and August 2024. Although extensive studies have been conducted on extreme heat (71 studies), drought (45 studies), extreme precipitation events (52 studies), and flooding (34 studies), important themes such as air quality (10 studies), chemical water quality (8 studies) and natural disasters (8) have been understudied. Additionally, this scoping review revealed a geographical gap in climate change and health studies, as only 24 out of 53 countries in sub-Saharan Africa were represented. The key deficiencies identified include limited funding, technological constraints, inadequate climate policies, and a lack of community-focused adaptation plans. Moreover, this review highlights the urgent need for resilient healthcare systems capable of addressing climate-related health risks effectively. Addressing these gaps is essential for developing targeted strategies to mitigate climate change’s health impacts and increase resilience in SSA communities. This review aims to inform policymakers, researchers, and stakeholders about critical areas requiring attention and investment by enhancing our understanding of these challenges and gaps. Strengthening research capacities, fostering collaboration, and implementing evidence-based policies are imperative steps toward achieving sustainable health outcomes in the face of a changing climate in sub-Saharan Africa.

## Introduction

Climate change poses a significant and escalating threat to public health globally, with particularly profound implications for vulnerable regions such as sub-Saharan Africa (SSA) and other developing countries. Characterized by diverse climatic conditions and socioeconomic challenges, SSA faces heightened susceptibility to the multifaceted impacts of climate change (1–7). Extreme heatwaves, shifting precipitation patterns, increased flooding frequency and intensity, and prolonged droughts directly influence health outcomes in the region (8–11). These environmental shifts exacerbate existing health disparities and introduce new risks, impacting the well-being of more than one billion people (12) in SSA.

In sub-Saharan Africa and other developing countries, the impact of climate change will be more dire due to unaddressed structural and socioeconomic vulnerabilities, poor governance, and inadequate supportive policies to address climate change and health issues. These unaddressed issues have resulted in other challenges, including water insecurity, poor healthcare infrastructure, a lag in research and training needed to address climate issues, and others (13–15). For instance, in places with a high burden of diarrhea disease due to a lack of Water, Sanitation, and Hygiene (WASH) services, climate change poses additional risks, as countries are now facing more diarrhea diseases and, in some areas, cholera outbreaks, most of which are attributed to climate change (2,16–20). Health conditions that are expected to be exacerbated in addition to waterborne and water-related diseases include foodborne diseases, vectorborne diseases, mental health, high morbidity and mortality due to heat stroke, malnutrition, and many others associated with natural disasters. Some countries are already experiencing higher rates than normal from malnutrition, diarrhea cholera, and vector-borne diseases such as dengue (21). In these countries, we are already seeing overwhelmed healthcare systems that are not equipped to address these public health challenges (14,22,23).

Although some African countries have developed plans for adaptation and mitigation strategies as required by the UN (24), there is little evidence on whether and how these plans are implemented. In addition, limited research has been conducted in SSA to guide country-specific plans for adaptation and mitigation and other associated measures to address or reduce the impact of climate change on health (25,26). Limited data is collected at the ground level to guide some of these plans and interventions. Most plans and research approaches are based on broader global perspectives (7,27–29). Given that SSA has unique challenges, research to guide adaptation, mitigation, and resilience efforts must be unique and specific to the region, if not country-specific. Given these challenges, SSA needs to scale up research, education, climate change financing, and policy – all collectively, to move forward toward sustainable solutions for long-term climate change resilience. Most importantly, there is a need to set a road map for priorities to guide efforts, focusing on addressing challenges specific to the region.

To understand the research that has been conducted to guide some of these efforts, this scoping review was conducted to examine current research on climate change and health impacts, policy, and to identify existing gaps in research and the need for policy-making to guide interventions to improve public health, climate change resilience in Africa. This scoping review aims to improve our understanding of where SSA is conducting research to generate specific data to address the complex challenges related to health outcomes due to climate change. The specific research questions guiding this review are as follows:

1. What are the geographic and thematic gaps in SSA’s climate change and health research, and how do these gaps affect our understanding of the region’s climate-induced health outcomes and vulnerabilities?
2. How do extreme weather events, such as heatwaves, droughts, and floods, interact with social determinants of health to influence health vulnerabilities and outcomes?
3. What practical solutions and community-based adaptation strategies can be developed and implemented to address air quality, water quality, and extreme heat, enhancing resilience and mitigating health impacts ?
4. How are climate change and health studies funded?

## Methods

A scoping review was conducted on PubMed, Embase, Web of Science and Scopus using the keywords “climate change,” “health,” and “sub-Saharan Africa” and imported to Covidence (30) to streamline and facilitate the review. The Covidence software follows the PRISMA approach for scoping review (Figure 1). Climate change impacts relevant to sub-Saharan Africa include extreme heat, food security, microbial and chemical water quality, flooding, and drought. A total of 7851 journal articles were identified using the keywords and inclusion criteria. No white papers, gray literature, review papers, or other sources were included. Papers were included if they (a) addressed the conceptualization of climate change in sub-Saharan Africa, (b) explored the impacts of climate change on health outcomes, (c) focused on relevant climate change impacts, including water quality, flooding, and drought and other health outcomes (d) were published between January 1, 2001, and August 1, 2024, and (d) were written in English. On the other hand, papers were excluded if they were not from the sub-Saharan Africa region or if they mentioned climate change impacts not relevant to the African context (e.g., changes in snowmelt or wildfires). Of the 7851 identified studies, 921 were duplicates, and 6628 were excluded because they did not meet the inclusion criteria. A total of 302 studies were screened for eligibility. Of the 302 studies, 149 were excluded; 53 were reviews, 2 were animal health research, 44 were malaria research, 23 were opinion pieces and correspondences, and 27 studies had the wrong geographical settings, study design or outcomes. In total, 153 studies were included for review (Figure 1). The screening of studies was conducted by two people, to minimize biases resulting from a single reviewer.

**Figure 1:**
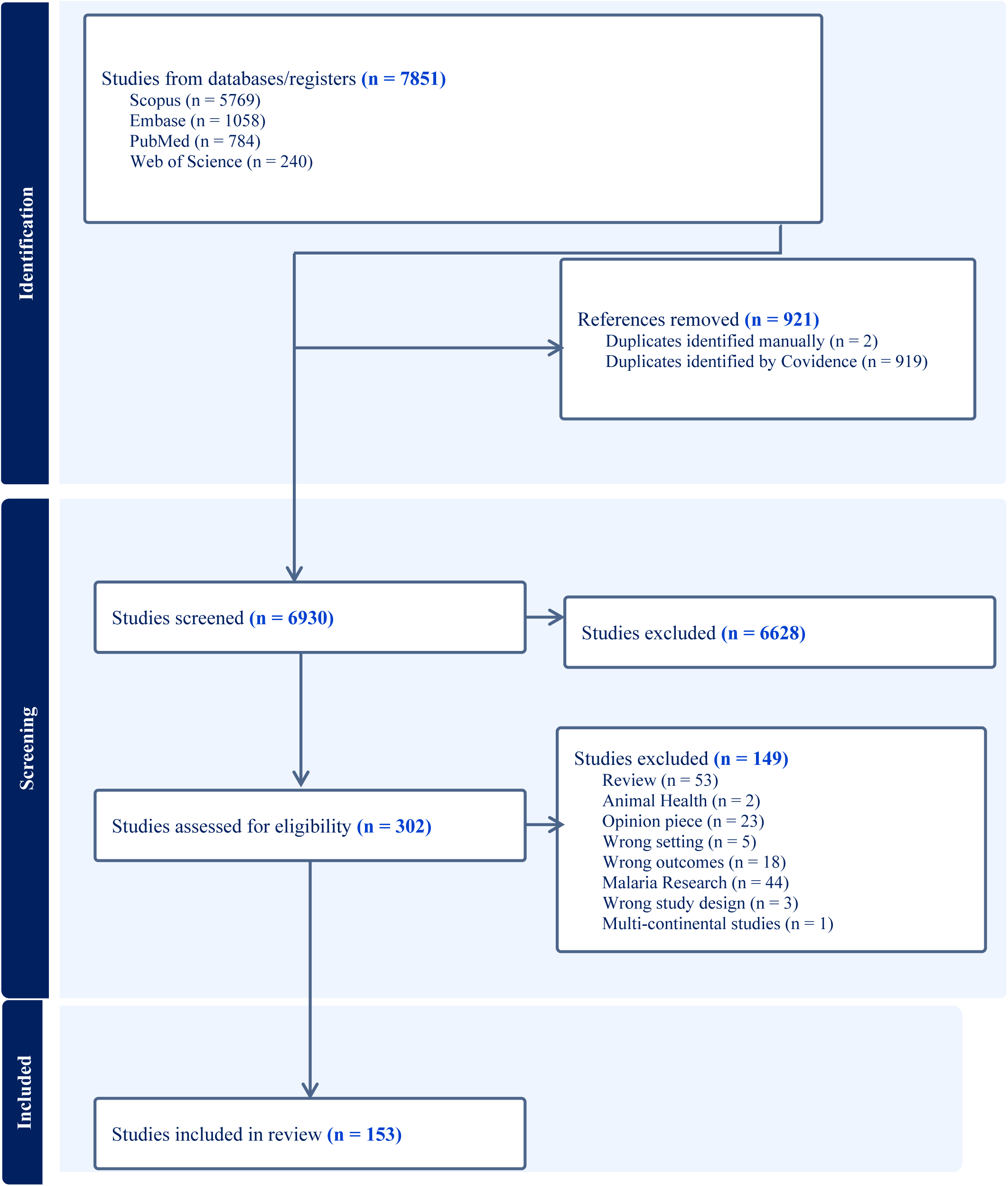
PRISMA flowchart showing the number of articles identified in PubMed, screened and included in the scoping review.

The included articles were thematically grouped to explore the various impacts of climate change in sub-Saharan Africa identified in the literature. Additionally, the literature was examined to identify the health outcomes associated with these impacts of climate change. The studies were further categorized thematically into the following groups: highlighting research needs, informing science, providing solutions and knowledge to combat climate impacts, highlighting policy needs, and identifying resource needs. Additionally, countries, where previous climate change and health studies were conducted, were visualized on a heatmap, providing insight into which countries have been studied extensively and where significant gaps in information exist. These thematic groupings enabled a comprehensive understanding of how climate change affects the region and its inhabitants. By categorizing the literature in this way, we could pinpoint specific gaps in research and policy that need to be addressed. The map further highlighted underresearched regions, emphasizing the need for more focused studies in those areas. Furthermore, the analysis helped to identify practical solutions and resources that can be mobilized to mitigate the adverse effects of climate change on health outcomes in sub-Saharan Africa. This approach ensures that the findings contribute to academic knowledge and provide actionable insights for policymakers, researchers, and practitioners working to combat climate change and its impacts on public health.

## Results

The majority of the studies published on climate change and health in sub-Saharan Africa between 2001 and August 2024 focused on extreme heat (71 studies), drought (45 studies), extreme precipitation events (52 studies) and flooding (34 studies). Additionally, a moderate number of studies have focused on infectious diseases (24 studies) and microbial water quality (23 studies). For almost 24 years, only 10 studies have been published on climate change and air quality (7 studies), chemical water quality (8) and natural disasters (8) (Table 1).

**Table 1.** Climate Change Impacts in sub-Saharan Africa Identified by Past Studies.

| Climate Impact | n | Studies |
| --- | --- | --- |
| Air Quality | 10 | (31–40) |
| Drought | 45 | (2,14,33,39,41–80) |
| Extreme Heat | 71 | (4,10,11,14,18,19,32,36,37,39,40,43,49,53,56,57,59–61,68–70,73,77,78,81–126) |
| Extreme Precipitation | 52 | (4,14,18,19,34,36,37,39,40,43,45,48,53,56–59,64,65,70,72,73,77,78,80,81,83,85,88,95,99,100,108–110,112,114,117,121,122,124–135) |
| Flooding | 34 | (2,2,4,8,14,19,40,45,53,57–61,64,65,68–71,73,77,80,83,87,88,98,99,121,126,130,136–139) |
| Microbial Water quality | 23 | (20,40,92,112,126,128,129,131,138,140–154) |
| Infectious Diseases | 23 | (1,11,14,72,73,91,92,122,131,141,146,154–164) |
| Chemical Water quality | 8 | (40,140,142,144,145,148,149,165) |
| Natural Disasters | 8 | (8,16,59,60,64,73,98,125) |

Of the 153 publications reviewed, 20 of the publications discussed and highlighted research needs, 58 studies informed science by providing insight into climate trends and impacts, and 15 studies highlighted policy; 43 publications discussed specific solutions and knowledge to help mitigate the impact of climate change on health and resource needs. Additionally, eleven studies explored multiple themes (Table 2).

**Table 2.** Research Categories and Number of Studies in Identified Categories from Previous Studies in Sub-Saharan Africa.

|  | n | References |
| --- | --- | --- |
| Highlight research needs | 20 | (19,35,45,54,62,67–69,71,86,88,90,91,94–96,103,106,127,132) |
| Informs science - Provides insight for climate trends and impacts | 58 | (1,4,8,11,33,37–39,43,51,56,60,70,72,74,75,79,98,99,102,104,105,108–116,119–122,124,129–131,133–136,139,146,148–150,152–154,158,162–164,166,167) |
| Provides solution and knowledge to combat climate impacts | 43 | (16,31,40,40,42,44,46,50,58,61,63–67,78,78,81,84,85,87,97,100,107,117,117,125,125,128,141–145,147,151,151,155,157,160,168–170) |
| Highlights policy needs | 15 | (14,34,53,57,59,93,101,118,123,137,156,159,171) |
| Highlights resources needs | 6 | (41,55,77,83,126,172) |
| Multiple Categories in a single study |  |  |
| Highlight research needs; Highlights policy needs | 3 | (32,92,161) |
| Highlights policy needs: Highlights resources needs | 1 | (20) |
| Informs science - Provides insight for climate trends and impacts; Highlights resources needs | 2 | (48,80) |
| Informs science - Provides insight for climate trends and impacts; Highlights policy | 2 | (73,76) |
| Provides solution and knowledge to combat climate impacts; Highlights policy needs | 1 | (2) |
| Provides solution and knowledge to combat climate impacts; Highlights policy needs; Highlights resources needs | 1 | (140) |
| Provides solution and knowledge to combat climate impacts; Informs the state of climate science in Africa | 1 | (36,47) |

With a particular focus on climate change and health, previous studies have focused on different health outcomes associated with a changing climate. Some of the major health outcomes included water, sanitation and hygiene issues (n=57), food security and malnutrition (n=40), physical illness (n=32) and health risks associated with pathogens (n=26). The remaining 53 articles highlighted the impact of climate change on health, specifically on health issues such as loss of livelihood due to natural disasters, climate induced displacement, mental health, gender-based violence, HIV and death (Table 3).

**Table 3.**
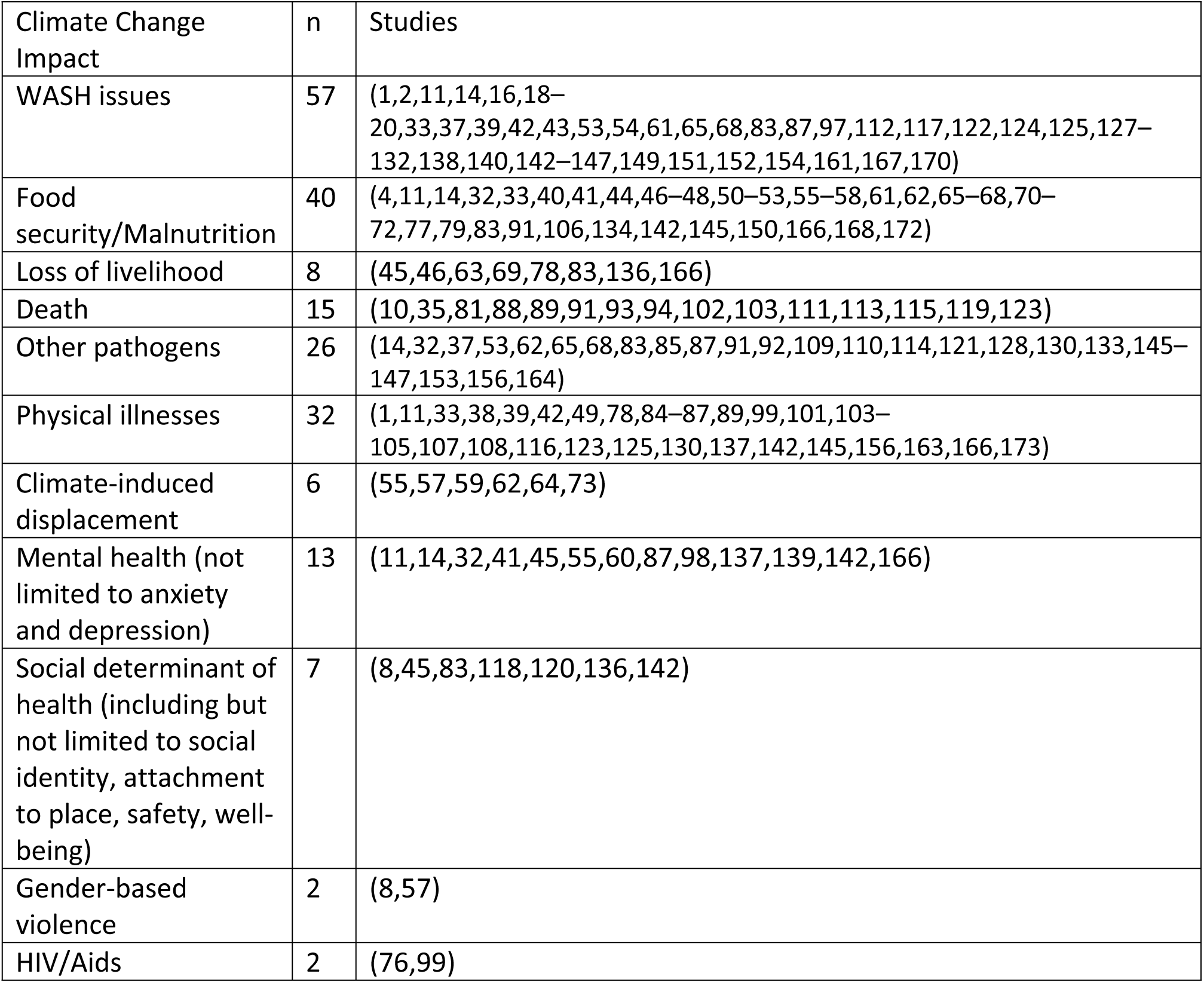
Climate Change and Health Issues in sub-Saharan Africa Identified by Past Studies.

This scoping review also examined specific countries in sub-Saharan Africa where climate change and health research originated. The findings were visualized on a map of Africa, with color coding from 4 to 24 studies, where a dark hue represents countries with more research and a lighter hue indicates countries with fewer studies (Figure 2). Countries shown in gray represent those with no published studies on climate change and health between 2001 and August 2024 that fit our criteria. The map shows that most of the studies were conducted in South and East Africa, with South Africa and Kenya having 24 and 22 published studies, respectively, followed by Ghana (14), Tanzania (10), and Burkina Faso (9). Twenty-six studies included in this study were large scale studies conducted across multiple SSA countries. Only 24 out of 53 countries in sub-Saharan Africa have published studies on climate change and health over the past 24 years and 15 countries had less than 5 published studies on climate change and human health (Supplemental Table 1).

**Figure 2.**
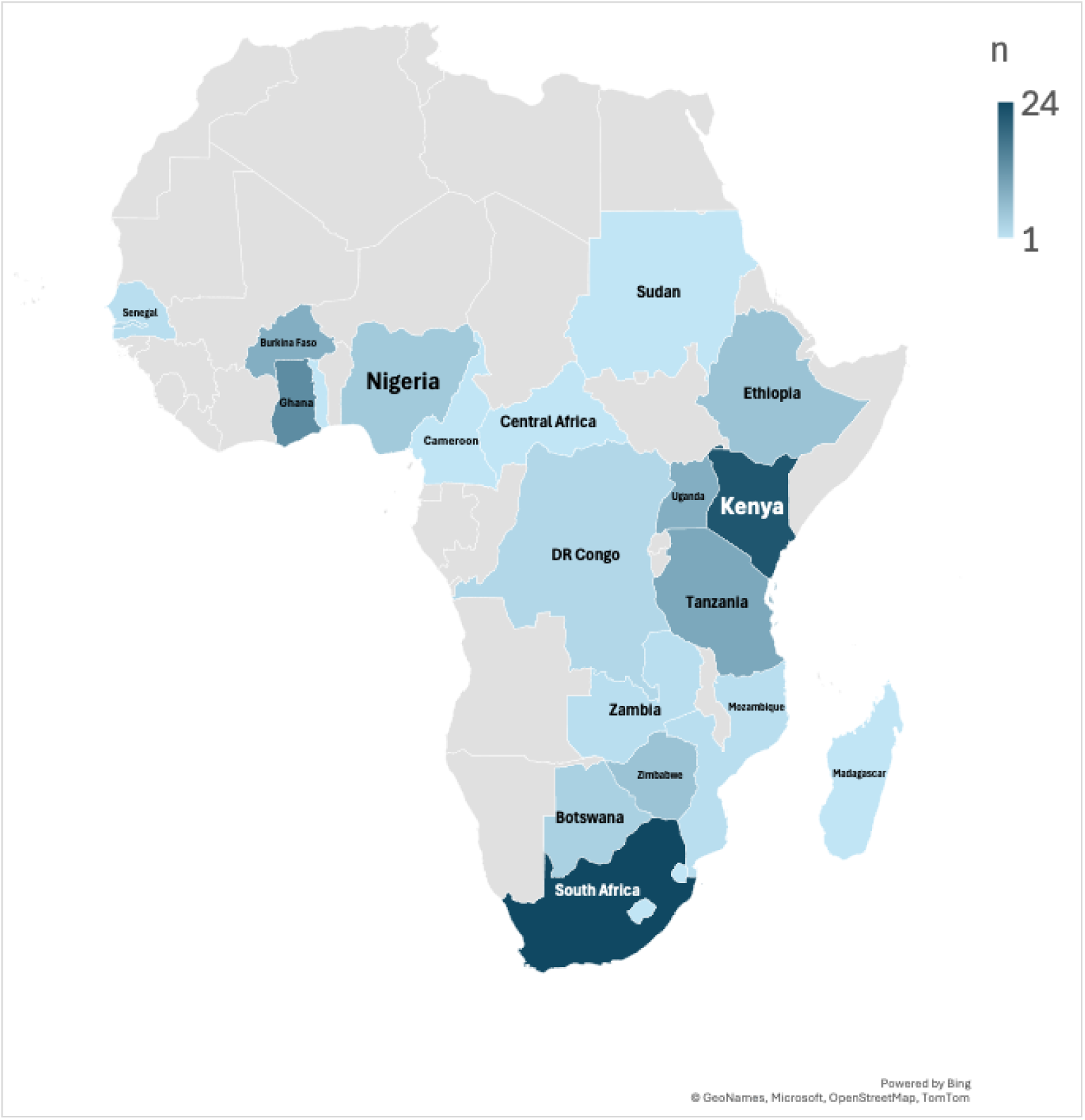
Map showing Number of Climate Change and Health Studies From Countries in sub-Saharan Africa Since 2001.

Research funding sources were also a focus of this study. Of the studies considered, 45% (n= 69) were funded by external grants, while 55% (n=83) were self-funded by researchers, declared as “no external funding” in research publication (Figure 3). Of the 45% (n= 69) of the research funded by external grants, 88.4% (n=61) were grants provided by international agencies across North America, Europe and Asia, including but not limited to DAAD, the Wellcome Trust, the Taiwan Ministry of Science and Technology Grant, the World Health Organization, the United States National Science Foundation, the Public Health Agency of Canada, the National Institutes of Health, the Rockefeller Foundation, the International Climate Change Information and Research Programme and Commonwealth. Funding was also provided by universities in the Global North, such as the University of Guelph. Only seven studies (11.6%) were funded through local funding provided by government agencies of sub-Saharan countries; four of those were provided by the South African Medical Research Council, one was a grant from Fundo Nacional de Investigação (FNI) in Mozambique, one was a grant from the Ethiopian Institute of Water Resources and one from the African Climate Change Fellowship Program (ACCFP).

**Figure 3.**
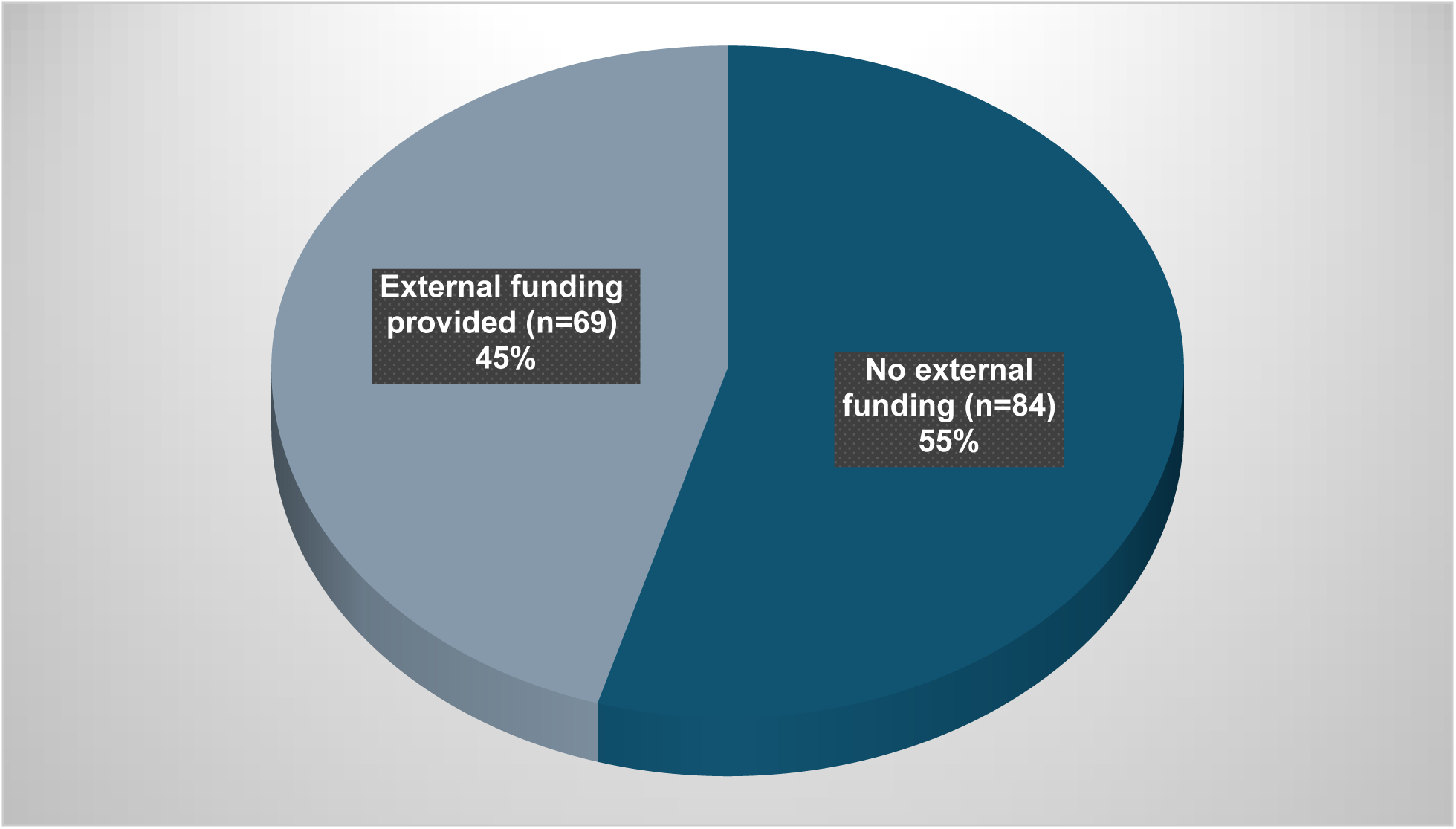
How are climate studies funded in sub-Saharan Africa?

Additionally, this study explored the impact of climate change on vulnerability and the health of vulnerable groups (Supplemental Figure 1). Among the studies reviewed, 38 did not consider vulnerability in their research. Of those who did, 29 focused exclusively on children, while 19 addressed children along with other vulnerable groups, such as elderly individuals, pregnant women, disabled individuals, low-income groups, and immunocompromised individuals. Two studies exclusively focused on immunocompromised groups, nine studies specifically examined gendered vulnerabilities, with eight focusing on women and one on men. Three studies investigated the impacts of climate change on the elderly population, and an additional two investigated the effects on impoverished populations. Three studies focused on pregnant women and two studies focused on mining workers. Finally, one study examined the impacts of climate change on the health of internally displaced populations and on local and indigenous communities.

This research also explored the various adaptation strategies proposed to mitigate climate change impacts on SSA, as suggested by the authors of the included studies. Thirty-seven studies (24%) had no proposed adaptation strategies. Adaptation strategies were grouped into themes including technology gaps, policy modifications, community-focused adaptation plans, climate preparedness programs and the need for funding. The majority of studies proposed a community-focused adaptation plan and the inclusion of local context in climate intervention (n=37, 24%) as the most effective adaptation strategy to help residents of SSA minimize the impacts of climate change. Improved technology was the second highest proposed adaptation strategy suggested by the studies reviewed (n=32, 21%), and the authors proposed technological improvements such as early warning systems, remote sensing, robust meteorological technologies, water treatment systems, vaccination, disease surveillance systems, energy-saving air conditioning, and climate-smart agricultural technologies. A total of 16% (n=25) of the studies discussed inadequate local funding as a significant barrier to climate change adaptation and proposed an increased budget for climate-related concerns. Similarly, an additional 16% (n=25) of the studies highlighted the need for climate preparedness programs among citizens, government officials and health workers. Finally, 13% (n=20) of the studies further describe policy modification, specifically suggesting holistic and concerted efforts by individuals, multiple government agencies working together and donor agencies in the fight against climate change and resulting health impacts.

## Discussion

The impact of climate change is already apparent in SSA, as the research reviewed in this study indicates that regions is already being affected by drought (33,47,63,91), extreme heat (9,84,88,90,94,166) and microbial and chemical water quality issues, leading to infectious diseases such as cholera and diarrhea outbreaks as a result of compounding climate extremes on water access and quality (9,16,87,138,143,149,174). Additionally, there have been reports of natural disasters, such as floods (8,68,137,149) and cyclones (16), all contributing to negative health outcomes in addition to economic costs.

The intensified climate events on the continent are leading to several physical, mental and emotional health impacts. Most of the studies in this review focused on WASH issues, food security, physical illnesses such as heat exhaustion, and pathogen infections such as cholera and malnutrition. However, the social, emotional and mental aspects of climate change have been relatively understudied. Loss of livelihood due to the impact of floods, extreme temperature and drought on agricultural, resource-based occupation and outdoor informal economies has been reported in a few studies in the sub-Saharan African context (83,136). Loss of livelihood is often intertwined with disrupted community ties, loss of identity, climate-induced displacement, climate anxiety, depression and gender-based violence (55,57,59,83,166). Therefore, climate change is leading to cascading outcomes affecting all aspects of health and wellness in the region. There is an urgent need for accelerated climate change research in Africa, home to more than 1 billion people and experiencing rapid population growth. Despite the critical need for solutions, only 43 of the publications reviewed in this study discussed specific strategies and knowledge to mitigate the impact of climate change on health. These publications included contributions to new research, evaluations of existing mitigation efforts, and insights into issues unique to the continent, such as lack of access to basic services and nutrition. Several studies have provided specific mitigations, considerations, and recommendations. For example, a study in Ethiopia examined childhood diarrhea, finding a high incidence at the beginning of dry seasons (43). Other studies have addressed climate-specific issues such as extreme temperatures, precipitation, and mortality and proposed mitigation steps to reduce hospital admissions, morbidity, and mortality (81,84,85), including within-hospital settings. The challenges related to water, sanitation, and hygiene (WASH) were also highlighted, with some studies identifying opportunities for integrating climate adaptation into WASH development planning (2). The findings and recommendations from these studies could be implemented in other regions facing similar challenges. Research on water burdens and gender inequalities has emphasized specific strategies for reducing these disparities (42). Overall, these studies offer valuable insights and recommendations that are specific to the region, that can be applied to mitigate the impact of climate change on health across SSA.

This review revealed existing gaps and opportunities in climate research in SSA. First, geographic disparities exist, with some countries being more studied than others. This disparity can lead to an incomplete understanding of what is needed for the continent or country-specific interventions. Even in countries where research has been conducted, authors have acknowledged the need for more primary climate data to promote data-driven decision-making. (14,71,171). Expanding research efforts to include understudied countries can provide a more comprehensive understanding of climate change and health impact across SSA, ensuring that attention is given to those most impacted by climate change and the opportunity to learn from each other.

Some considerations need to be considered when discussing climate change solutions for SSA. For instance, we need to integrate and discuss social determinants of health as we think of climate change and health. Only five studies highlighted or discussed the associations between health, social determinants of health, and climate change (8,45,83,136,142). However, it is well understood that the work a person does, where people live, their educational level, and income level all play a role in determining health (175). More research is needed to account for climate change as a determinant of health, especially since developing countries will be impacted the most. Additionally, there are only a few studies on how climate change leads to gender-based violence. However, relationships between climate change, loss of livelihood, feelings of emasculation and gender-based violence have been reported in sub-Saharan Africa (8,57). Many studies have not adequately considered the vulnerability of different population groups with an intersectional lens. Although a good number of studies mentioned vulnerable groups such as children, pregnant women and elderly people, most of the reviewed papers mentioned only vulnerability in passing, and 38 studies did not consider vulnerability at all in their research. More comprehensive research is needed to focus on community heterogeneity and intersectionality among the most affected groups. Therefore, while the physical impact of climate change has been relatively well studied in certain countries of sub-Saharan Africa, there is a need for more research on the interconnections between climate change and socioeconomic wellness indicators.

Further, while some climate themes have been extensively studied, others have been less studied and require further attention in sub-Saharan Africa. Of the 302 published articles assessed for eligibility, 44 were specific to malaria research. This was one of the largest groups for research, followed by WASH (57). Rightfully, malaria research has been ongoing for a long time and is well-established (176). However, there is a lack of research in other areas of climate change and health, such as extreme heat, natural disasters, and air pollution. For instance, only 10 studies highlighted research related to air pollution (31–37) and two specifically related to climate change and cardiovascular diseases (34,42). This is a significant finding, mainly since the Lancet Global Burden of Disease (2016) reported that 33% of the global burden of stroke is attributed to environmental factors such as air pollution and lead exposure (177). This study also revealed that air pollution has emerged as a significant contributor to the global burden of stroke in low- and middle-income countries (LMIC) (177). Climate change is expected to increase drought and air pollution in some regions of the world, including SSA (21). Despite this significance, research in this area is almost nonexistent and requires urgent attention.

A few other studies offer concrete pathways and recommendations on what countries need to focus on and prioritize to reduce the impact of climate change. Technological advancement was one of the most common emerging themes from the included studies, although implementing the suggested technologies might be far-fetched in SSA landscapes. For example, in areas where infectious diseases are a concern, authors recommend focusing on early warning signs, disease forecasting, and climate disaster mitigation (14,71) to strengthen public health systems to cope with and reduce climate change impacts. However, it is difficult in developing countries to focus on early warning systems or disease forecasting when basic solutions for preventing diseases, such as access to safe and clean water or sanitation, are still lacking.

For instance, without the additional burden of intensified climate impacts, current water and sanitation infrastructure or healthcare infrastructure is inadequate to protect public health (2,17,19,127,143). Some countries are already experiencing preventable waterborne diseases that could be addressed by having adequate water and sanitation infrastructure. Some examples include the cholera outbreak in Tanzania in 2023, ongoing in 2024, and another in June 2024 in Nigeria (16,18,178–180), both attributed to climate change. In addition, lack of technologies and inadequate electricity coverage create barriers to implementing other simple solutions to combat extreme heat conditions (166) and a lack of improved agricultural technologies to combat food insecurity(4). For example, irrigation systems are creating vulnerabilities further intensified by climate change. The continent must catch up to address current poor living conditions and, at the same time, accelerate the research needed to move to the 21^st^ century to address climate change issues.

In tandem with inadequate climate policies, a lack of funding was also identified as limiting climate change research progress in SSA. In the present study, more than half of the studies reviewed were self-funded by researchers. The large amount of self-funded research highlights the financial constraints researchers face, which can limit the scope, scale, and depth of their studies, ultimately hindering the development of comprehensive climate change mitigation and adaptation strategies. Additionally, only seven of the 153 selected studies were funded by local governmental agencies, revealing a much deeper systemic issue about the perceived relevance of climate change impacts in this region. Therefore, there is an urgent need for increased investments in climate research (2,19,20,41,65,67,83). Funding climate research will help inform climate decision-making and policies (71). There is also a lack of climate consideration in government policies (2). To ensure that climate investment and increased funding reap the maximum benefits, there is a need for multiagency collaboration between government agencies to inform policies that achieve economic, environmental, social and health justice (45,173). Local residents and external donor agencies should also be included in climate strategizing to ensure maximum benefits (99). Addressing these limitations through enhanced funding, comprehensive policies, and inclusive collaboration will be pivotal for effectively mitigating the impacts of climate change in SSA.

Even with limited resources, countries can strategize and prioritize actions towards climate change. One thing is evident: the issue is not a lack of understanding of the urgency to address climate change. Studies evaluating climate change perception in various African countries have shown that most of the population is familiar with climate change and have experienced it to some exten(14,59,63,64,67,181). However, there is insufficient data due to technological and resource limitations to provide tailored interventions at the community and national, and lack of national policies. For instance, in Zimbabwe, following the devastation caused by Cyclone Ida, natural disasters exposed another problem—food insecurity. A study of 19 health facilities conducted after Cyclone Ida revealed that 94% of the facilities were not equipped to address malnutrition, either due to lack of proper training, inadequate staffing, or failure to follow proper protocols(66). With climate change, areas with limited resources will be most affected, and there is a need for adequate systems to address climate change and health issues. Therefore, community-focused adaptation plans are necessary, taking into account the strengths and vulnerabilities of individual communities and countries. These details can only be revealed through community-level or national research, which is often obscured by large-resolution, aggregated datasets produced by major development agencies.

There is also a dire need to build capacity for health sector personnel through climate preparedness programs that integrate climate resilience into public health systems. Educational and research institutions can offer expertise in addressing regional or national climate change solutions. These programs should focus on enhancing the ability of healthcare infrastructure to withstand climate-related events and training healthcare professionals to respond effectively to climate-induced health issues (6,21,53,64,130,150,182). Additionally, community-based interventions are essential for educating and empowering local populations on climate-related health risks and adaptive practices. Incorporating climate education in formal and informal places of learning in communities, has shown to increase ability to identify vulnerability and build adaptive capacity (53,60). These climate education programs may include utilizing traditional knowledge to inform strategies to reduce individual- and community-level vulnerabilities, improve infrastructure, create multiple climate-resilient income streams, and address the intersectional vulnerabilities that climate change exposes (60). Due to the large population of young people in Africa, strategies targeting young people are paramount (59). Educational programs may also target improved monitoring and surveillance systems (14,171) to fill the data gap and improve data-driven decision-making. These efforts will ensure that public health systems are resilient, adaptive, and equipped to protect the well-being of all individuals, especially the most vulnerable populations.

## Conclusion and Recommendations

The scoping review of climate change and health research in sub-Saharan Africa from 2001 to August 2024 reveals significant gaps and opportunities. Despite increasing recognition of climate change impacts, the region remains underresearched, especially in areas beyond extreme heat, drought, precipitation events, air quality, and flooding. This has left substantial gaps in understanding the full spectrum of climate-induced health outcomes and the intersectional vulnerabilities that various populations face. The reviewed studies predominantly originated from South and East Africa, with South Africa, Kenya, and Ghana being the relatively most studied countries. This leaves many nations with little to no climate health research, highlighting the urgent need for more geographically diverse studies to develop a comprehensive understanding of climate impacts in specific countries across the region. The limited funding and support for climate change research in sub-Saharan Africa, with a significant reliance on self-funded studies, further exacerbates these challenges. Increased investment from both international and local sources is essential to support robust research initiatives. Additionally, the insufficient focus on the social determinants of health and the mental, emotional, and social aspects of climate change calls for a broader research agenda that integrates these critical dimensions.

Among other recommendations, and in terms of competing resources and disasters, SSA needs to have a clear map and priorities to build climate change resilience. There is a need to 1) conduct research that is relevant to the continent to provide long-term solutions to address climate changes, especially in water, air, and extreme heat; 2) accelerate building robust water and energy infrastructure to bring the continent to the 21^st^ century to help build climate change resilience; 3) build resilient health systems and implement training programs for healthcare professionals; 4) accelerate knowledge by building capacity and greening higher education to produce leaders capable of understanding and providing solutions that are specific to their regions; and 5) increase funding from both international donors and local governments to support comprehensive climate and health research. Investments should focus on underresearched countries and diverse climate health topics to ensure a holistic understanding of climate impacts across the region; 6) community-based adaptation strategies should be promoted, incorporating local knowledge and contexts to enhance resilience and adaptive capacity at the grassroots level; and 7) a focus should be placed on vulnerable populations to ensure that the specific needs and vulnerabilities of these groups are addressed; and 8) monitoring and surveillance systems should be improved through investments in technological advancements.

In the short term, climate change needs to be framed as a threat to health and economic development for the continent and, therefore, integrated into health policy and planning, and other economic planning and budgets to facilitate funding for climate change actions. Collaboration between governments, research institutions, and international agencies is necessary to foster holistic policies addressing environmental, social, and health justice. Enhancing the capacity of educational and research institutions to address regional and national climate change solutions is crucial for equipping future generations to address climate challenges. Research that addresses practical solutions for air quality, water quality, and extreme heat should be prioritized, as these areas are critical for mitigating health impacts and improving overall resilience to climate change.

## Data Availability

All relevant data are within the manuscript and its Supporting Information files.

## Aknowledgement

None, only authors were involves in the scoping review.

## Author contributions

Questions and Conceptualization: A.K, G. C., V. O., Review of Journal Articles: A.K, G. C., V. O., Writing of Original Draft Preparation: A.K., Writing-Review & Editing: A.K, G. C., V. O., M. H., Supervision: A.K

## Declaration of Competing Interest

The authors declare that they have no known competing financial interests that could have appeared to influence the work reported in this paper.

## Funding

No external funding

## Appendices

**Figure 1.**
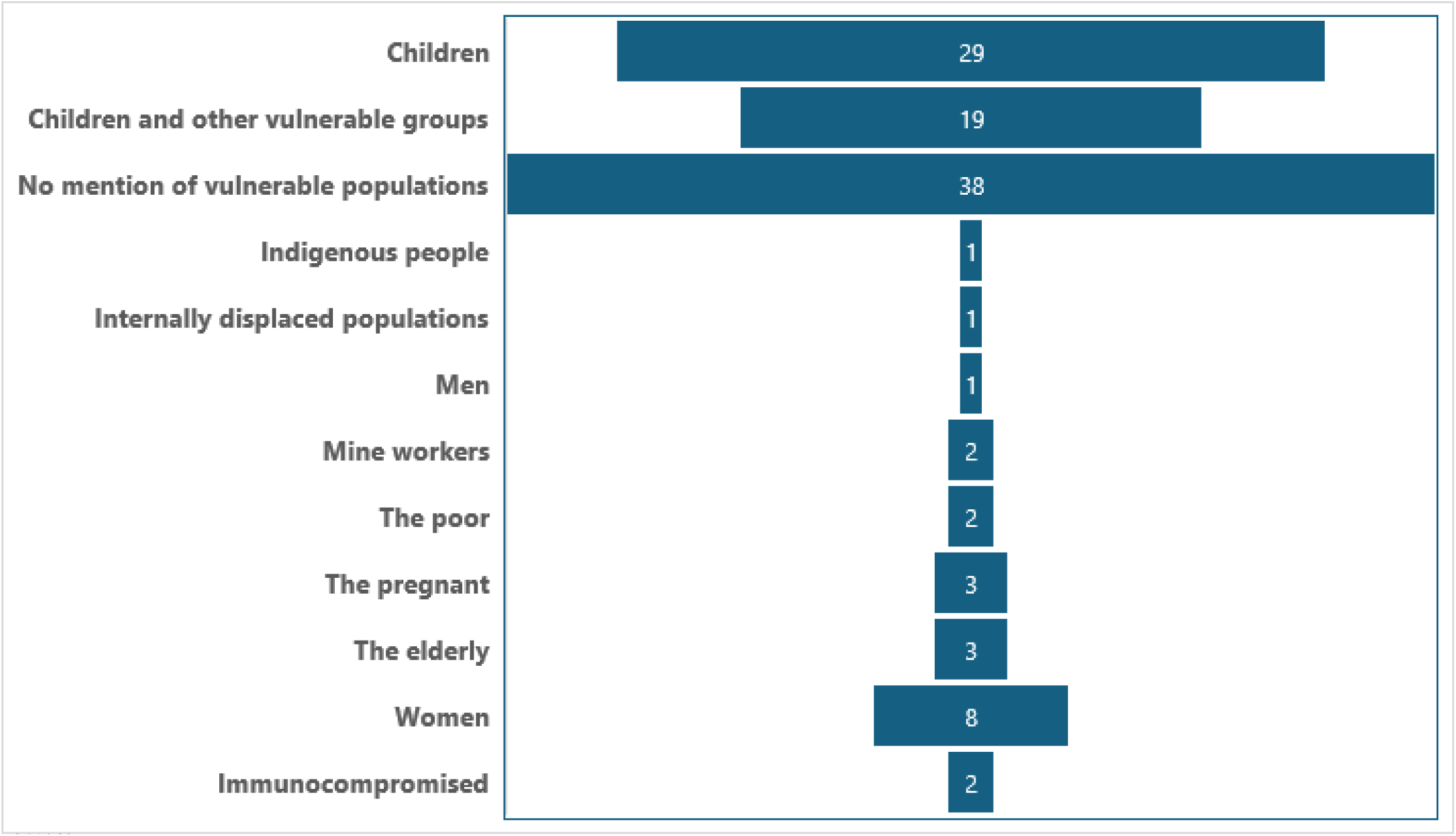
Vulnerable Groups Identified in Past Studies and the Number of times the group was mentioned.

**Table 1.** Number of Studies on Climate Change and Health by Country in Sub-Saharan Africa.

| Country | n | Studies |
| --- | --- | --- |
| Burkina Faso | 9 | (67,68,81,88,95,121,134,157,166) |
| Botswana | 4 | (1,138) |
| Kenya | 22 | (8,38,45,46,48,53,80,97,98,104,108,109,112,140,142,147,150,153,156,160,163,172) |
| South Africa | 24 | (4,10,14,18,32,34,36,37,40,65,84,93,94,107,110,111,115,124,139,143,148,163,167,170) |
| Mozambique | 2 | (16,129) |
| Tanzania | 10 | (20,35,60,100,103,109,116,128,152,159) |
| Uganda | 9 | (33,42,47,70,83,85,109,116,118) |
| Ghana | 14 | (2,49,59,72,87,90,92,101,102,123,136,137,141,173) |
| Nigeria | 5 | (39,77,117,130,151) |
| Cameroon | 1 | (131) |
| Zimbabwe | 6 | (66,69,96,113,125,158) |
| Ethiopia | 6 | (43,51,62,79,149,169) |
| DR Congo | 3 | (99,126,164) |
| Gambia | 3 | (50,86,105) |
| Eswatini | 1 | (44) |
| Zambia | 2 | (63,154) |
| Madagascar | 1 | (61) |
| Togo | 1 | (31) |
| Central Africa | 1 | (89) |
| Sudan | 1 | (114) |
| Senegal | 2 | (132,135) |
| Lesotho | 1 | (76) |
| Malawi | 1 | (74) |
| Rwanda | 1 | (78) |
| Study across multiple countries in SSA | 26 | (11,19,52,55,56,58,73,75,82,89,91,106,119,120,127,133,144–146,155,161,162,168,171) |

## Notes

### Competing Interest Statement

The authors have declared no competing interest.

### Funding Statement

The author(s) received no specific funding for this work.

